# An Indicator Cell Assay-based Multivariate Blood Test for Early Detection of Alzheimer’s Disease

**DOI:** 10.1101/2025.09.15.25335782

**Authors:** Yijun Sherry Qi, Leslie Rae Miller, Mark D. D‘Ascenzo, Jason D. Berndt, G. Adam Whitney, Fergal Duffy, Samuel A. Danziger, Elaine Peskind, Ge Li, Colin L. Masters, Christopher Fowler, Australian Imaging Biomarkers and Lifestyle Study of Ageing (AIBL) Research Group, Robert Lipshutz, John D. Aitchison, Jennifer J. Smith

## Abstract

The indicator cell assay platform (iCAP) is a novel next-generation approach for blood-based diagnostics that uses standardized cells as biosensors to amplify weak disease signals in blood. We developed an Alzheimer’s disease iCAP (AD-iCAP) for early detection at the mild cognitive impairment/mild dementia stages. To develop the assay, patient plasma is incubated with standardized neurons, which transduce complex circulating signals into gene-expression readouts used to train multivariate disease classifiers via machine learning. We applied systems biology analyses (e.g., GSEA, PCA, correlation/network analyses) to optimize analytical and computational parameters, and then evaluated a locked model in a study with retrospectively collected samples. Performance was AUC 0.64 (95% CI 0.51–0.78, n=82) on an independent external-validation set and AUC 0.77 (95% CI 0.57–0.96, n=23) on a blind set, supporting prospective confirmation in a larger cohort. To overcome pre-analytical noise and reduce bias in feature-selection, modeling was done using a fixed panel of 84 candidate genes chosen a priori from an external AD-iCAP dataset generated with 5XFAD mouse plasma. Despite using no AD-specific prior knowledge in this approach, the assay readout was enriched for Alzheimer’s-relevant pathways, including cholesterol biosynthesis, synaptic structure/neurotransmission and PIK3/AKT activation. Because the assay senses a multivalent cellular response, which is orthogonal to circulating amyloid or tau measurements, AD-iCAP may complement existing blood tests, and its multivariate readout offers a path to precision-medicine applications such as patient stratification for treatment response.

## Introduction

Alzheimer’s disease (AD) is a multifaceted disorder characterized by complex mechanisms involving amyloid plaques, tau tangles, neuroinflammation, oxidative stress and various other pathological mechanisms that contribute to cognitive decline. It is ranked as the 6th leading cause of death in the United States, affecting 1 in 9 people aged over 65, equivalent to approximately 6.9 million individuals,^1^ and the associated annual healthcare cost is estimated at $360 billion.^1^ There are no treatments to prevent or cure AD and until recently, there were also no effective disease modifying treatments.^2,3^ The progression of AD drugs through clinical phases and regulatory review faces considerable challenges, with success rates being the lowest for any therapeutic area.^4^

Failures in clinical trials have been attributed in part to targeting advanced disease stages, and the disease’s inherent complexity and heterogeneity^3^. Recent breakthroughs have marked a departure from this trend; two new amyloid antibody drugs, Lecanemab and Donanemab, have had unprecedented success in phase 3 clinical trials focusing on the mild cognitive impairment (MCI) and early AD stages by significantly reducing cognitive decline by 27%^5^ and 36%^6^ compared with placebo, respectively. These successes mark the first significant drug efficacy and the validation of amyloid as a contributor to AD; however, there are concerns about safety risks, high costs and modest clinical benefits. Treatments are needed that address patient heterogeneity and target the complex mechanisms of AD beyond amyloid pathology.^3^

Cost-effective, multivariate blood tests are needed for the early detection of AD to enable early treatment and to stratify patients for clinical trial selection.^7^ Blood tests for plasma (amyloid-beta 42/amyloid beta 40) (Aβ42/Aβ40), p-tau181 and p-tau217 have been developed that have high concordance with amyloid measurements in the brain by PET.^8–12^ However, to support development of more effective treatments for AD, multivariate blood tests are needed that can stratify patients based on varying response to treatments, underlying pathological mechanisms, and/or disease trajectories among individuals. Such tests could be developed to characterize treatment efficacy on distinct patient subsets, which could detect clinical effectiveness for abandoned drugs and new drugs in development. They may also be useful for risk prediction models capable of estimating various risk levels through continuous assay readouts, contributing significantly to informed decision-making.^13^

Development of reliable multivariate blood-based diagnostics for patient stratification is impeded by low abundance of circulating biomarkers and high levels of biological noise.^14–16^ To address these barriers, we developed the Indicator Cell Assay Platform (iCAP), which uses standardized cultured cells as biosensors to detect and amplify disease signals in patient blood.^17^ This cell-based approach offers several conceptual advantages: broad responsiveness to diverse inputs, signal amplification, standardized cellular context for normalization, and multivariate gene expression outputs well-suited for classifying heterogeneous diseases. These features position iCAP as a novel platform for developing generalizable, machine learning–based diagnostics.

We have developed an iCAP for early detection of AD (AD-iCAP) at the stage of mild cognitive impairment (MCI)/mild dementia. Developing the test involved exposing cultured standardized neurons to plasma from case and control patients, measuring the gene expression response of the neurons and using it to develop multivariate models to predict the disease class of the patients (Fig. 1). Building on a prior proof-of-concept study that established feasibility of the assay using a limited set of samples,^17^ we conducted a clinical validation study in which we optimized assay and modeling parameters and trained a new classifier using samples from three independent sources. Feature selection was independent of the study samples and prior knowledge of AD biology, potentially improving robustness and generalizability of the model, and yielding new biomarkers that might contribute to a deeper understanding of disease mechanisms. The development of this multivariate model for early Alzheimer’s disease diagnosis establishes a strong foundation for a prospective confirmation in a larger cohort and advancing the platform’s use in patient stratification for drug development and other precision medicine applications.

**Figure 1.**
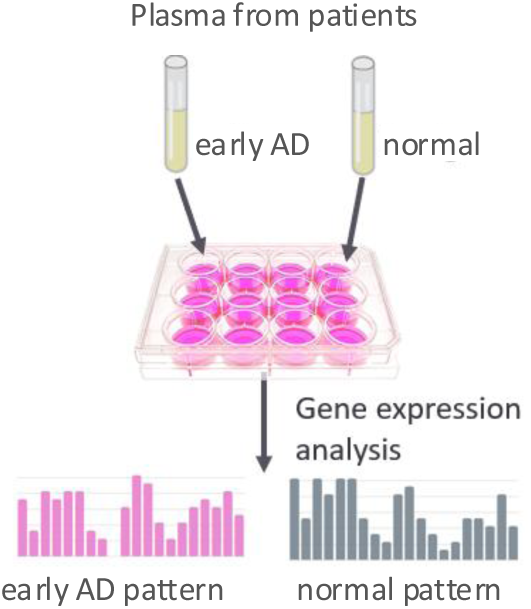
The iCAP for blood-based diagnostics. Standardized cells are exposed to plasma from patients. Gene expression readout of cells is used to develop machine learning-based models to predict disease. The iCAP transforms weak, variable blood signals into a stable, system-level gene expression readout, unlocking the power of systems biology for blood-based diagnostics

## Materials and Methods

In accordance with the guidelines for transparent reporting, we followed the TRIPOD (Transparent Reporting of a multivariable prediction model for individual Prognosis or Diagnosis) guidelines in the Methods and Results sections of this report.^18^

### Participants

This retrospective study used archived human plasma samples and associated clinical data previously collected from participants in other observational studies (described below). The WCG IRB determined that the study does not constitute human subjects research. There were three cohorts of samples from three different sites. All procedures were approved by the institutional review boards (IRBs) of each study site, and all participants provided written informed consent. The study consent forms had provisions allowing use of their samples for future research purposes. Patient identifiable information was not provided to the research team and was not used in this study. Plasma samples were from four classes of participants defined in Table I and Data file 1. Diagnoses were made by physicians based on neuropsychological testing and clinical and laboratory evaluations using criteria established by a National Institute of Neurological and Communicative Disorders and Stroke (NINCDS) and the Alzheimer’s Disease and Related

**Table I.** Plasma sample characteristics and cohorts used for AD-iCAP development. Cohorts 1, 2, and 3 correspond to AIBL (Florey Institute of Neuroscience and Mental Health), ADRC Washington University, and ADRC University of Washington, respectively. Samples from each preAD, MCI and normal class were acquired from each source in roughly 1:1:1 ratio. Numbers in brackets show cohort size after sample exclusion in Fig. 3. An additional 46 nonAD dementia samples were included from cohort 1 for exploration. Normal and preAD patient had CDR scores of 0. For MCI patients, 64% had mild cognitive impairment (CDR 0.5), 34% had mild dementia (CDR 1) and 2% had moderate dementia (CDR 2). Samples from cohorts 2-3 were stored at -80℃, whereas samples from cohort 1 were collected in the presence of a plasmalogen stabilizer and stored in liquid nitrogen before commencement of the study. Aβ, amyloid-β.

| Class | Description | Sample number |  |  |
| --- | --- | --- | --- | --- |
|  |  | cohort 1 | cohort 2 | cohort 3 |
|  |  | Florey | Wash U | VA |
| preAD | Preclinical AD patients: cognitively normal with Aβ pathology and no MCI or AD diagnosis | 44 (19) | 25 (22) | 50 (35) |
| MCI | Early symptomatic AD patients: cognitive impairment, Aβ pathology and may have MCI or AD diagnosis | 66 (46) | 24 (19) | 53 (35) |
| Normal | Healthy age-matched controls: cognitively normal, no Aβ pathology and no diagnosis of MCI or AD | 87 (63) | 25 (21) | 50 (32) |
| nonAD | Patients with cognitive impairment and no Aβ pathology and no diagnosis of AD | 46 (27) | (1) | 0 |
| subtotal |  | 243 (155) | 74 (63) | 153 (102) |
| total |  | 470 (320) |  |  |

Disorders Association (ADRDA) workgroup in 1984^19^. Level of cognitive impairment was measured using Clinical Dementia Rating (CDR).^20^ Amyloid-β pathology was measured by CSF amyloid-β (1–42) protein fragment (Aβ42) levels or by neuroimaging with amyloid-β tracers (PiB-PET) with additional CSF Aβ42 levels provided for some patients. Samples were from three different cohorts and for each cohort, there were differences in how amyloid-β pathology was measured as well as the sample filters applied to improve confidence of disease classes and omit patients in transition between classes.

**Cohort 1** samples were from participants enrolled in Australian Imaging, Biomarkers and Lifestyle Study of Ageing (AIBL) described previously^21^ (http://aibl.csiro.au/), with oversight and approval from the institutional ethics committees of Austin Health, St. Vincent’s Health, Hollywood Private Hospital and Edith Cowan University. The longitudinal study was initiated in 2006 to test various biomarkers, cognitive parameters, and lifestyle factors for association with AD development. All volunteers gave their written informed consent prior to participating in the study. The study consisted of 1,166 participants over the age of 60 years who were analyzed longitudinally at 18-month intervals for blood draws and for clinical assessments. Disease assessment at each timepoint was performed by a clinical review panel to improve consistency using the NINCDS-ADRDA international work group criteria for AD diagnosis^22^. Amyloid-β pathology was assessed primarily by PET scan (with 11^C^-PiB, ^18^F-florbetapir or ^18^F-flutemetamol) with or without supporting CSF Aβ42 analysis. Samples for this study consisted of samples collected between 2007-2017 with a balanced number of classes from each of two different collection sites, Melbourne and Perth.

**Cohort 2** samples were from participants enrolled in studies of normal aging and dementia at the Knight ADRC at Washington University in Saint Louis collected between (2009-2014). Participants were community-dwelling volunteers (age 45-84) enrolled in longitudinal studies of normal aging and dementia. For this cohort, all amyloid-β pathology was assessed by measuring CSF Aβ42 levels using INNO-BIA ALZBIO3 test. Given known assay drift over time regarding the Aβ42 value that best corresponds with amyloid positivity, a conservative cut-off was used for assigning Aβ42 negativity (>1000 pg/mL) and positivity (<500 pg/mL) to increase the odds of samples being correctly assigned respective amyloid categories. Therefore, there were no samples in the cohort with Aβ42 values between 500-1000 pg/mL. Other exclusion criteria were presence of neurological, psychiatric, or systemic medical illness that might compromise longitudinal study participation, or medical contraindication to lumbar puncture (LP) for CSF collection or PiB-PET.

**Cohort 3** samples were from University of Washington Veterans Affairs (VA) Puget Sound Health Care System Alzheimer’s Disease Research Center (ADRC) study related to memory loss with oversight and approval by the IRBs of the University of Washington. Samples were collected between 2002 and 2008 from community-dwelling volunteers ages 51-87. For this cohort, amyloid-β pathology was assessed by measuring CSF Aβ42 levels using multi-analyte Luminex assay (INNO-BIA AlzBio3) with positive and negative pathology corresponding to a previously established threshold of ≤ 192 pg/mL, and > 192 pg/mL, respectively.^23^ CSF collection and analysis were as described previously.^24^ Samples classified as normal were also CSF Tau negative to avoid inclusion of suspected non-AD pathogenesis (SNAP). Other exclusion criteria were neurological or psychiatric disorders that could affect cognitive function, history of moderate to severe head injury, unstable major medical conditions, and use of illegal drugs or stimulants.

### Human Plasma characteristics

For all three cohorts, whole blood was obtained from each subject by venipuncture after overnight fasting and before lumbar puncture. For cohorts 2 and 3, plasma was prepared by centrifugation at 2,000 x *g* for 15 minutes at 4°C in EDTA tubes, then aliquoted and frozen at 84°C or lower. For cohort 1, plasma samples were prepared the same way except two measures were implemented to improve plasma quality: 1) EDTA tubes contained prostaglandin E1 (Sapphire Biosciences, 33.3 ng/ml), a plasmalogen stabilizer to reduced platelet activation and contamination of plasma during plasma preparation and storage, and 2) samples were stored in liquid nitrogen instead of at -80°C after collection and before shipping to PreCyte for analysis to improve preservation of plasma biomarkers. iCAP assays were performed on plasma samples thawed once after initial freezing.

### Experimental Study design

The experimental study design including assay development (gray), and clinical validation (pink) is summarized in Figure 2. The patient samples, cohorts, classes and clinical characteristics used for defining classes are in Data file 1 and Table I. The study included retrospective analysis of 470 human plasma samples. These samples included subjects at two stages of AD progression (preclinical AD (preAD), and MCI) and unaffected control samples. Samples were from three different cohorts, each with a ∼1:1:1 ratio of samples from each preAD, MCI and normal class, with gender and age were roughly balanced between classes. In addition, for exploratory purposes, a limited number of samples from a non-AD dementia class were included and grouped with the normal class in some models. Samples were processed in 26 experimental batches and 7 RNA-seq batches. Before processing, a portion of the samples were assigned to a blind test set for final testing. All samples were blind to scientist performing the experiments and the test set was also blind to data scientists until after final predictions were made. To reduce bias, each partition had equivalent class ratios. Each iCAP batch contained 22 participant samples from the same cohort with a roughly balanced number of samples from each class and each partition. For quality control, 2 reference controls were included in each iCAP batch, which were aliquots of a plasma sample from a healthy male donor at age 40. All data were collected over the course of 10 months. Before modeling, various data filters were applied to improve sample and data quality (see Sample Exclusion and Results Sections).

**Figure 2.**
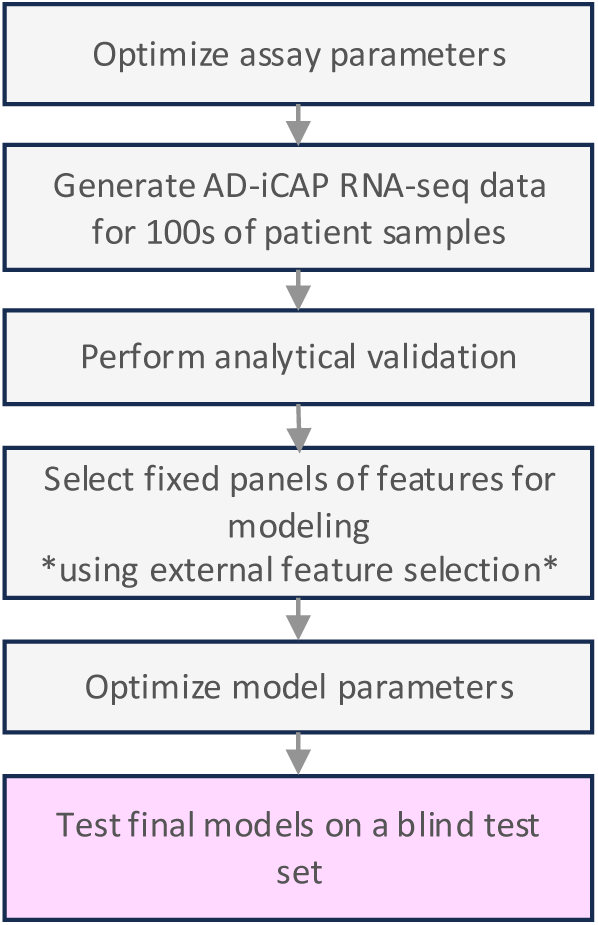
Stages of developing the AD-iCAP to predict early-stage AD from plasma samples.

### Analytical parameters of the AD-iCAP assay

Unless otherwise stated, AD-iCAP experiments were performed using the standard conditions described below. Indicator cells were a commercially available, pan-neuronal population of glutamatergic and GABAergic neurons derived from induced pluripotent stem cells (iPSCs) (iCell Neurons from Fujifilm (formerly Cellular Dynamics International (CDI)). Cells were grown according to manufacturer’s instructions with a modification to the culture duration described below. 12-well Eppendorf moat plates were coated with 0.01% poly-L-ornithine solution (Sigma P4957) for 1 h followed by 3.3 µg/mL laminin (Sigma L2020) for 1 h. Next, iCell neurons were thawed and plated a density of 488,875 cells/well in a total volume of 1 mL/well. Medium was exchanged 24 h after plating. After culturing for a total of 48 h, cells were exposed to 5% plasma containing 0.2 mg/mL heparin sodium salt (Stem Cell Technologies, 07980) in PBS for 6 h. For experiments with human plasma samples, RNA was isolated manually using an RNeasy Micro kit (Qiagen) and for experiments with mouse plasma samples, automated RNA isolation was performed using a MagMax mirVana kit (Invitrogen, A27828) on either a KingFisher Flex or a Kingfisher Duo Prime liquid handling robot as per the manufacturer’s recommendation (Thermo Fisher Scientific). Total RNA was stored at -80°C.

### AD-iCAP gene expression studies using RNA-seq

Unless otherwise stated, 100 - 250 ng of total RNA from each iCAP sample was used for automated library preparation and RNA sequencing (RNA-seq) performed by Azenta Life Sciences (South Plainfield, NJ, USA) (formerly Genewiz). Strand-specific library prep was performed with PolyA selection using TruSeq RNA Library Prep kit (Illumina) with unique dual indices (IDT) and resulting DNA samples were pooled and spread evenly across lanes and sequenced on a HiSeq 4000 (Illumina) with paired-end 150 bp reads yielding 20-30 M total reads per sample. RNAseq data were processed using a custom workflow using SnakeMake workflow management system for reproducibility.^25^ The workflow included adapter read trimming using Trimmomatic,^26^ genome reference alignment to GRCh37 using STAR,^27^ and gene-level transcript quantification using R-featureCounts.^28^ Read duplicates were removed by using –ignoreDups setting in R:featureCounts. Quality was assessed using MultiQC, dupRadar and GATK.^29^

RNAseq data from all 26 AD-iCAP batches were merged into a single file and several preprocessing steps were performed in R to detect and correct technical noise in the data to reduce error as described below. Genes were filtered to remove those below the limit of detection. To do this, ERCC controls were removed and a histogram of expression levels for each gene across all patient samples was generated, showing a bimodal curve indicating some genes were below the limit of detection (Fig. S2*A*). A threshold was selected to separate the curves (red dashed line) corresponding to 35 counts and genes were filtered to remove those below this threshold for greater than 10% of the samples. Data were normalized for heteroskedasticity by variance stabilizing transformation (VST) using the R-DESeq2 package and for inter-iCAP batch variation using removeBatchEffect from the R-limma package.^30^ Counts were adjusted to correct for GC bias using either full quantile normalization (FQN)^31^ or conditional quantile normalization (CQN)^32^ determined by the R-EdgeR package.^33^ Differential expression analysis was performed using R-DESeq2.^34^ GSEA analysis was applied using the R-fgsea package.

### Development of AD-iCAP standard controls

Assay standards were developed to monitor performance during development and deployment of the assay. To monitor the reproducibility of the gene expression readout, a biological standard was developed consisting of human plasma from an unaffected male of 40 years of age that is applied the indicator cells and processed alongside patient samples.

### Stimulating the AD-iCAP with pathogenic Aβ

We measured the response of the indicator cells to pathogenic versus non-pathogenic Aβ. First, we generated two peptides: 1) oligomerized Aβ25-35 (oAβ25-35), the minimal pathogenic fragment of Aβ (GSNKGAIIGLM), and 2) scrambled Aβ35-25 (sAβ35-25), a non-oligomerizing sequence of amino acids in Aβ25-35 in scrambled order (MAKGINGISGL). Lyophilized peptides oAβ25-35 and sAβ35-25 (AnaSpec, AS-24227 and AS-61971, respectively) were made to 2.5 mM in deionized water, and stored at -80°C in 50 µL aliquots. Aliquots were thawed and incubated at room temperature for 3 hours for complete oligomerization of the pathogenic peptide^35^ and applied to indicator cells at 5-10 µM final concentration and incubated 6 h or 24 h as indicated.

To identify a differential readout for monitoring assay performance, three technical replicates each of oAβ25-35 versus sAβ35-25 were analyzed in the AD-iCAP using standard conditions except neurons were pre-cultured for 2 weeks. RNA was isolated and analyzed by nCounter with Human Neuropathology Panel v1.0 consisting of 770 genes following manufacturer’s recommendations (analyzed by the Genomics Core of Fred Hutchison Cancer Research Center). Data were normalized to housekeeping genes in nSolver and genes with mean counts < 30 were filtered out. Differential expression analyses were performed using R-limma. Processed data are in Data file 2.

### Mouse plasma for cross sectional AD-iCAP study

The mice used for this study were from Jackson Laboratory colony 006554 (genetic background B6SJLF1/J; strain B6SJL-Tg (APPSwFlLon,PSEN1*M146L*L286V) 6799Vas/Mmjax). Hemizygous mice from this colony overexpress mutant human Aβ (A4) precursor protein 695 (APP) with the Swedish (K670N, M671L), Florida (I716V), and London (V717I) Familial AD (FAD) mutations along with human presenilin 1 (PS1) harboring two FAD mutations, M146L and L286V and develop AD symptoms in an age dependent manner. The plasma samples for this study were collected from female hemizygous mice at 12, 21 and 29 weeks of age, along with control plasma from gender- and age-matched non-carrier mice from the same colony by Jackson Laboratories under their breeding Services IACUC Protocol #03001. During aging, mice were housed at 3 mice per cage maximum. For EDTA plasma collection, mice were euthanized by CO_2_ asphyxiation following AVMA guidelines for the Euthanasia of Animals and the blood was collected via an open chest cardiac puncture. A new sterile needle and syringe were used for each mouse. Blood plasma was placed in an appropriate centrifuge tube and kept at room temperature for 20-30 minutes before being centrifuged. For each experimental group, plasma from 10 mice was pooled, aliquoted, flash frozen, stored at -80°C, and shipped on dry ice.

### Cross-sectional AD-iCAP study with mouse plasma

Pooled plasma samples from affected and non-carrier 5XFAD mice were analyzed in the AD-iCAP in a cross-sectional study. This was done to identify AD-specific differentially expressed genes to use as features for developing disease classification models with human AD-iCAP data. We analyzed plasma from mice at three ages (12, 21 and 28 weeks) corresponding to 3 disease stages in humans (preAD and two MCI stages) based on previous studies measuring cognitive decline^36^ and Aβ42 pathology in these mice.^37^ The experiment was repeated 3 times using three different pre-culture times for the neuron indicator cells (2, 5 and 14 days). For each of the 9 conditions, 4 technical replicates of pooled plasma from hemizygous and non-carrier mice were analyzed. 1µg of RNA from each AD-iCAP well was used for gene expression analysis by RNA-seq and data were processed using both CQN and FQN GC bias correction. Log_2_ fold change differential expression and false discovery rates were assessed for each condition using R-Limma/voom using both CQN and FQN corrected data for a total of 18 disease versus normal comparisons. Genes with FDR < 0.05 were considered to be significant.

### Sample size

Study sample size was based on balance of cost and power. An initial proof of concept study determined AD-iCAP to have an AUC of 0.64 (p-value 2.9E-2) before optimization. From this it was estimated that 30 samples per class in a validation set would be sufficient to detect significant AUC (with lower limit of 95% CI >0.5). Sample size was selected to have at least 30 samples of each class per each data partition.

### Sample exclusion

Sample exclusions were made: 1) to improve RNA-seq data quality or RNA integrity quality, and 2) to improve uniformity of sample characteristics between the cohorts. This ‘cohort harmonization’ specifically addressed a bimodal distribution in CSF Aβ levels in cohort 2 caused by site-specific inclusion criteria to improve class confidence. All three cohorts in the study had the same selection criteria (Table II), however there were differences in how Aβ42 pathology was measured (PET for cohort 1 versus CSF for cohorts 2 and 3) and in how the samples were prefiltered; for cohort 2 only, samples with CSF Aβ42 measurements close to the threshold were excluded from the repository to improve class confidence and distinction between the classes (see participants section). Therefore, to improve sample homogeneity, we made exclusions from cohorts 1 and 3 to similarly improve class distinction. For cohort 3, samples were excluded that had CSF Aβ42 levels that were close to the threshold of 192 pg/mL (see Data file 1). For cohort 1, 18 samples were excluded from subjects with discordant presence of Aβ42 pathology from PET versus CSF data at the time of blood draw, and 13 samples were excluded from subjects that did not have PET data at the time of blood draw and whose Aβ42 pathology could not be confidently ascertained from longitudinal data. These class confidence filters reduced diagnostic ambiguity, ensured cleaner class separation and improved similarity between the cohorts. All filtering was blind to sample and partition labels.

**Table II.**
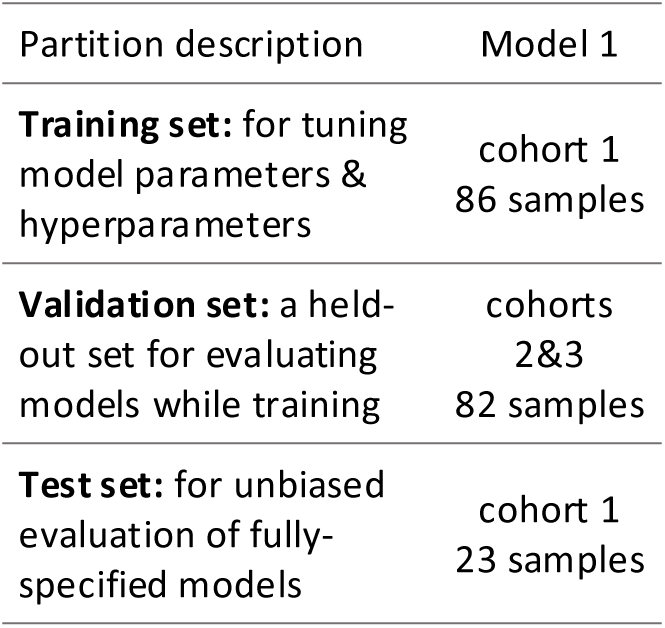
Sample partition for AD-iCAP Model M1.

**Table III.** Plasma sample characteristics across data partitions for Model 1. A strong covariate shift was observed between the training/test sets and the validation set, particularly with respect to sample storage duration. Additionally, Florey samples (used for the training and test sets) were processed with a plasmalogen stabilizer and stored in liquid nitrogen, whereas VA and WashU samples (used in the validation set) lacked stabilization and were stored at –80°C.

| Variable | N | data partition |  |  |  | p-value<sup>2</sup> |
| --- | --- | --- | --- | --- | --- | --- |
|  |  | Overall, N = 191<sup>1</sup> | train, N = 86<sup>1</sup> | validation, N = 82<sup>1</sup> | test, N = 23<sup>1</sup> |  |
| **status** | 191 |  |  |  |  | 0.41 |
| MCI |  | 87 (46%) | 38 (44%) | 41 (50%) | 8 (35%) |  |
| normal |  | 104 (54%) | 48 (56%) | 41 (50%) | 15 (65%) |  |
| **source** | 191 |  |  |  |  | <0.001 |
| Florey |  | 109 (57%) | 86 (100%) | 0 (0%) | 23 (100%) |  |
| VA |  | 52 (27%) | 0 (0%) | 52 (63%) | 0 (0%) |  |
| WashU |  | 30 (16%) | 0 (0%) | 30 (37%) | 0 (0%) |  |
| **gender** | 191 |  |  |  |  | 0.67 |
| female |  | 109 (57%) | 52 (60%) | 45 (55%) | 12 (52%) |  |
| male |  | 82 (43%) | 34 (40%) | 37 (45%) | 11 (48%) |  |
| **storage.yr** | 190 | 10.0 (9.0, 14.0) | 9.0 (7.0, 9.8) | 15.0 (11.0, 17.0) | 9.5 (8.0, 10.0) | <0.001 |
| Unknown |  | 1 | 0 | 0 | 1 |  |
| **apoe4** | 190 |  |  |  |  | 0.031 |
| 0 |  | 112 (59%) | 57 (66%) | 40 (49%) | 15 (65%) |  |
| 1 |  | 61 (32%) | 25 (29%) | 28 (35%) | 8 (35%) |  |
| 2 |  | 17 (8.9%) | 4 (4.7%) | 13 (16%) | 0 (0%) |  |
| Unknown |  | 1 | 0 | 1 | 0 |  |
| **age** | 188 | 72 (67, 78) | 74 (70, 78) | 69 (62, 77) | 72 (70, 77) | 0.002 |
| Unknown |  | 3 | 2 | 0 | 1 |  |

<sup>1</sup> Median (IQR) or Frequency (%)
<sup>2</sup> Pearson's Chi-squared test; Fisher's exact test; Kruskal-Wallis rank sum test

### Modeling with patient AD-iCAP data

#### Feature selection

Before disease modeling, the AD-iCAP RNA-seq gene expression data were filtered from ∼20,000 to 398 or 84 genes using independent data generated from the cross-sectional AD-iCAP study with mouse plasma. Two gene sets were used for modeling: Gene Set 1, 398 genes with significant differential expression in affected versus non-carrier mice in at least 1 of the 18 datasets, and Gene Set 2 , 84 genes with significant differential expression in at least 3 of the 18 datasets.

#### Sample partitions

Before any data generation or AD-iCAP analysis, samples from cohorts 1-3 were partitioned into training, validation and blind test sets defined in Table II, each with a 1:1:1 class ratio (preAD:MCI:normal). For this modeling study, the training and blind test sets were comprised of cohort 1 samples and the validation set was comprised of cohorts 2 and 3 (Table II). This partitioning was because cohort 1 had the largest sample size (Table I) and highest sample quality (see Participants section). Four class configurations were used for modeling: (1) MCI vs. normal, (2) preAD and MCI vs. normal and non-AD dementia, (3) preAD vs. normal, and (4) MCI vs. preAD, normal, and non-AD dementia.

#### Model development

Random Forest (RF) models were trained and validated. We used the RF classifier implemented in the randomForest package,^38^ accessed via the caret interface in R^39^ using the sample configurations and feature selection methods described above. Prior to model training, feature dimensionality was reduced using a two-step process: first, a correlation filter from the R-caret package (with a threshold of 0.8) was applied to remove highly correlated features, followed by Boruta feature reduction (to retain confirmed or tentative features) using the R-Boruta package.^40^ Mean Decrease Accuracy was used to assess feature importance during Boruta selection. In total, 240 models were developed, each with 500 trees based on combinations of the following factors: two feature sets, two mtry thresholds (ceiling and floor), four class comparisons, three feature reduction approaches (two with a correlation filter combined with Boruta feature reduction and one with no feature reduction) and five random seeds per model. Model performances were assessed by 5-fold cross-validation on the training set (using only left-out folds), and by predicting on the external validation set.

#### Models advanced to blind independent testing

Model M1, trained to classify MCI versus normal samples was advanced to blind-set evaluation based on two pre-specified criteria: 1) Discrimination on development data: significant performance on both repeated, stratified cross-validation of the training set and external validation (on cohorts 2–3), defined as an ROC AUC whose 95% CI lay entirely above 0.5; and 2) Adequate power: the blind test set had sufficient sample size to detect an AUC significantly >0.5 per our a priori power analysis.^41^.

Two other pre-specified models (M2 and M3 for classifying MCI versus normal) were evaluated in parallel with M1 on the same blind set. Their development process was identical except they were trained/validated on cohort 1 only (no external validation). Because M1 uniquely demonstrated transportability on independent cohorts, we designate M1 as the primary analysis. Performance of M1 on the blind set was evaluated by AUC of ROC with DeLong 95% CI, which was pre-specified as the primary endpoint. Control for family wise error rate was done using the Holm test. Although AUC is invariant to prevalence,^42^ we also calculated MCC as a secondary metric because of mild class imbalance in the blind test set; MCC significance was assessed via label-permutation on the blind set (10,000 permutations) with 95% CIs by bootstrap.

#### Other modeling

Before the current study using *external* feature selection, an earlier analysis was conducted using *internal* feature selection based on the training set. In that study, 12 fully parameterized models were tested on the blind test set across two validation attempts (each with parallel evaluation). This was conducted independently, and neither the test data nor the results were shared with the data scientists involved in the current study. In addition, pre-defined sample omission criteria (based on technical failures and patient metadata inconsistencies) were the same for both modeling studies.

### Authentication of Key resources

The same lot of iCell® neurons were used for all data generation used for modeling (lot 1225418). They had a certificate of analysis from Fujifilm (Madison, WI) and were guaranteed by morphology assessment and were determined to be 98% pure by staining with β-III tubulin (+)/ Nestin (-). β-III tubulin is a classic neuronal element of the tubulin family found almost exclusively in neurons but excludes other cell types of brain origin like glial cells, etc. Nestin is a type VI intermediate filament that is typically expressed in neuronal precursor cells but becomes downregulated upon differentiation. iCell® neurons are cortical neurons and are estimated to be 5% glutamatergic and 95% GABA neurons. The cells were guaranteed free of microbial contaminants and mycoplasma. Supplements and maintenance media were guaranteed free of microbial contaminants with low endotoxin contamination at <= 2EU/mL.

### Control for errors and bias

To reduce potential for biases, our sample workflow incorporated sample order randomization and class balancing at each step. In addition, samples were coded and researchers performing the AD-iCAP were blind to disease status and clinical data for all samples and data scientists were blinded to the classes and clinical data of the test set. For AD-iCAP cell culture, moat plates were used to reduce edge effect and middle wells were used for assay controls. Each AD-iCAP plate contained two control wells of either standard reference serum, which were used to monitor assay reproducibility and develop normalization approaches.

To control for bias and error in computational modeling, feature selection was done with external mouse data without using prior knowledge of Alzheimer’s biology. In addition, final models were validated with two sets of independent samples, including an external validation set and a blind test set. Validation with independent samples has been shown to control for overfitting and minimize overestimate of model performance, demonstrating generalizability despite sample size variations.^43^ All statistical analyses controlled for family-wise error rates.

### Data and software availability

All data needed to evaluate the conclusions in the paper are present in the paper and/or the Supplementary Materials. RNA-seq data related to this paper will be available from GEO under the accession numbers GSExxxx after publication. R code for disease classification model M1 will be available at http://doi.org/10.5281/zenodo.XXXXXX after publication.

## Results

We have developed the iCAP for blood-based diagnostics, a novel assay that uses cultured cells as biosensors. It capitalizes on the evolved ability of cells to detect weak signals in noisy environments to overcome low signal to noise ratio of blood biomarkers.^17^ To develop an assay, cultured indicator cells are exposed to patient plasma samples and the gene expression readout of the cells is measured and used to develop machine learning models to predict disease in new subjects (Fig. 1).

The goal of this study was to develop and validate an iCAP for early detection of AD (AD-iCAP). Our approach was to distinguish patients with early-stage AD (preAD and/or MCI) from age-matched controls using retrospective plasma from three sources (Table I). We optimized assay parameters, generated data from hundreds of subjects, and trained models using a rigorous two-step approach including model tuning using training and external validation sets, followed by blinded testing of selected locked-down models using an independent blind test set (Fig. 2). To reduce overfitting, gene expression features were selected externally from longitudinal AD-iCAP data generated with plasma from the 5XFAD mouse model. We followed TRIPOD and STARD reporting guidelines in the Methods and Results sections.^18,44^

### Assay Development

#### Optimization of AD-iCAP parameters

Initial AD-iCAP parameters were previously established in a proof-of-concept study and included using iCell® neurons as indicator cells after 5-days of pre-culturing, followed by 24-hour exposure to 10% patient plasma.^17^ As an initial step in this study, we performed an experimental parameterization analysis by analyzing technical replicates of patient samples under multiple experimental conditions with the goal of increasing the number of case versus control differentially expressed genes and improving indicator cell culture consistency. As a result, we reduced the pre-culture time to 2 days and reduced plasma exposure to 5% for 6 hours. These changes shortened assay time, improved RNA yield, and enhanced commercial viability of the assay (Fig. S1).

After optimization, assay reproducibility was assessed by measuring the expression of 182 genes across technical replicates of patient samples in the AD-iCAP using NanoString. The genes were selected as those with high biological variability (i.e. those with significant case versus control differential expression in sample subsets). The median coefficient of variation (CV) was ∼3.3%, with the highest variability observed for low-abundance transcripts (Fig. S4A).

#### Generation of AD-iCAP RNA-seq data from patient plasma samples

Patient plasma samples from four diagnostic groups (pre-MCI, MCI, normal, and non-AD dementia) were sourced from three cohorts (Table I). A total of 470 samples were partitioned into training, validation, and blind test sets and assayed using the optimized AD-iCAP parameters to generate total RNA for further processing by RNA-seq. Each batch included 22 patient samples with balanced classes and two technical replicates of a reference control sample. Class labels were hidden from technicians performing the AD-iCAP during sample processing and class labels of the test set remained blind to computational model developers until after model finalization.

Strand-specific RNA-seq was performed across seven batches using a HiSeq 4000 with unique dual indices and automated library prep. Subsequent data processing focused on detecting and correcting sources of analytical noise, which was essential for detecting early-stage disease signals. Technical noise was mitigated in three steps: (1) A lower limit of detection (LLD) was set, retaining 13,744 genes (Fig. S2A), (2) library size variability was corrected using VST (Fig. S2B– D), and (3) GC bias from library prep was corrected using FQN or CQN (Fig. S3A–D). Notably, GC bias correction appeared to reduce a technical artifact where variability of GC enrichment otherwise introduced bias between classes (e.g. compare Fig S3B before (left) and after correction (right). Although this variability might not substantially affect the outcomes of typical laboratory experiments with large effect sizes, our data highlights the necessity of robust preprocessing strategies in biomarker studies with subtle biological signals.

#### AD-iCAP analytical validation

Our analysis of within-batch AD-iCAP reproducibility showed low analytical variation of the cell-based assay (Fig. S4*A*). Next, we examined the aggregate RNA-seq dataset, including both patient samples and concurrently processed technical replicates of control samples, to determine whether biological variation could reliably be detected above analytical noise in a large-scale, multi-batch study. To do this, CVs for each gene were calculated across patient samples of multiple classes and multiple batches (capturing biological variation) and across technical replicates (capturing analytical variation) and compared. Generally, higher variability was observed across biological versus technical replicates, suggesting that biological variation is detectable above analytical noise (Fig. S4*B*). 250 genes were selected with a high ratio of biological to analytical variation (*∼2:1*, *blue points* in Fig. S4*B*) and used for principal component analysis (PCA) to further analyze the source of variation. Analysis of the 250 genes revealed that sample source had a strong influence on variation whereas case versus control differential expression was not observed (Fig. S4*C*).

In conclusion, analytical validation revealed low technical noise, with a median CV of approximately 3%. This finding was further supported by our analysis of the aggregate AD-iCAP dataset, which confirmed that patient-to-patient variability was detectable above the level of technical noise. However, further stratification of the data by PCA showed that disease-specific differential expression in AD was not detected, and that variability in gene expression was primarily due to sample source. This low signal-to-noise was likely due to pre-analytical noise from patient heterogeneity and/or site differences in sample collection or storage, issues well known in blood biomarker studies.^45,46^

#### External feature selection for modeling using a mouse model of AD

To overcome the preanalytical noise detected above and identify AD-iCAP features for modeling, we implemented external feature selection using AD-iCAP profiles generated from plasma of 5XFAD transgenic mice.^37^

The 5XFAD model harbors five AD-linked human mutations and hemizygous mice develop AD pathology in an age-dependent manner.^47^ We conducted a study of differential expression in the AD-iCAP using plasma from hemizygous mice versus non carrier littermates. We minimized noise from genetic, environmental, disease stage and technical sources by using age- and gender-matched littermates and pooling plasma from 10 mice per condition and running 4 technical replicates of each per class.

Longitudinal AD-iCAP data were generated from mice at 3, 5, and 7 months, spanning pre-symptomatic and MCI stages,^36,37^ under nine assay conditions (3 mouse ages x 3 neuron pre-culture times). After RNA-seq and data processing using two different GC bias correction methods, we obtained 18 differential expression datasets. From these data, we derived two feature sets: 398 genes detected in ≥1 dataset (Set 1) and 84 genes detected in ≥3 datasets (Set 2), all with adjusted p < 0.05. These two feature sets were used as fixed gene panels for subsequent model development.

This external feature selection approach effectively reduced noise, enabling detection of disease-specific signals and generation of two fixed gene feature sets for modeling. The use of fixed feature sets in multivariate model development has been successfully applied in other gene expression models and is critical for robust model performance.^48,49^ Selecting features independently of the study samples and without reliance on prior knowledge of AD biology further minimizes bias and error in modeling. Moreover, establishing such AD-related panels lays the groundwork for regulatory approval, standardized clinical deployment, and the development of AD-iCAPs for AD patient stratification.

#### Training AD-iCAP models for predicting early-stage AD

The human plasma AD-iCAP data were used to train classification models using the fixed 398- and 84-gene sets derived from mouse AD-iCAP profiles (Set 1 and Set 2). Before modeling, we first applied exclusion criteria to the AD-iCAP data to improve data quality and class clarity (Fig. 3; Data file 1). Next, we defined the modeling partition. Because cohort 1 was identified as the highest quality (see Methods, Table I), models were trained on cohort 1 and validated against cohorts 2 and 3, and final blind testing was performed using cohort 1 (Table II). Importantly, this partitioning was done without altering the *a priori* partitioning of the blind samples.

**Figure 3.**
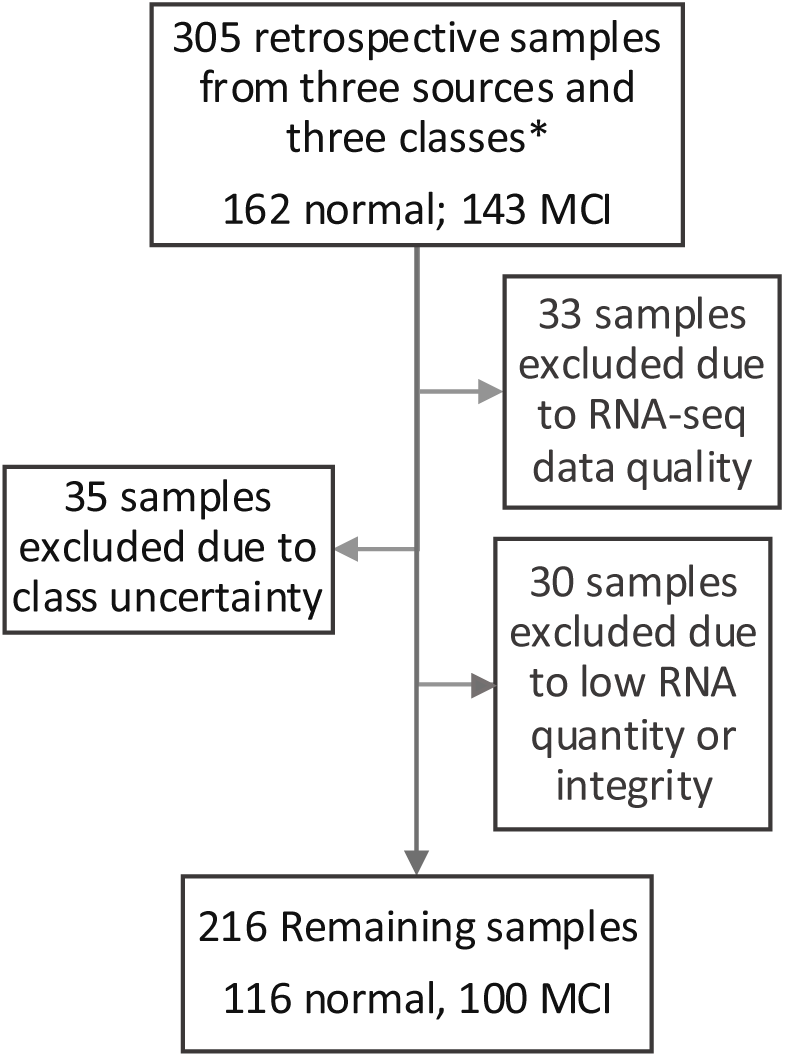
Sample exclusion flow chart for Model 1. A total of 89 samples were excluded due to either low class confidence (based on patient metadata) or analytical failures. Samples with class uncertainty included those with conflicting Aβ pathology from cerebrospinal fluid (CSF) and PET imaging, CSF Aβ₄₂ values near the diagnostic threshold, or absence of Aβ data at the time of blood draw. Aβ, amyloid beta; CSF, cerebrospinal fluid; PET, positron emission tomography.

Models were developed in R using the randomForest package implemented in the caret package^39^ using Boruta-based feature reduction.^40^ 48 model configurations outlined in the methods section were trained each with five random seeds. Performance was evaluated on held out folds in 5-fold cross-validation using the training set followed by validation on the external validation set.

### Assay validation

#### Testing fully parameterized models on a held-out blind set

From the 240 models, we selected a model (M1) for blind testing that met two pre-specified criteria: (1) significant performance on both training and external validation sets based on AUC of ROC, and (2) sufficient power to detect significance of performance on the blind set, as determined by power calculations.^41^

Model M1 achieved an AUC of 0.64 (95% CI: 0.51–0.76) on the external validation set in the parameterization study and AUC of 0.77 (95% CI: 0.57–0.96) in final blind testing on the test set (Fig. 4; Data file 3; MCC = 0.456 (permutation p = 0.032)). The external validation performance was lower than blind-set performance. This is likely because the validation set (cohorts 2 and 3) comprised externally sourced samples with less ideal pre-analytical storage compared with the other partitions (cohort 1) (see methods), indicating a covariate shift between the validation set versus the training and blind sets.

**Figure 4.**
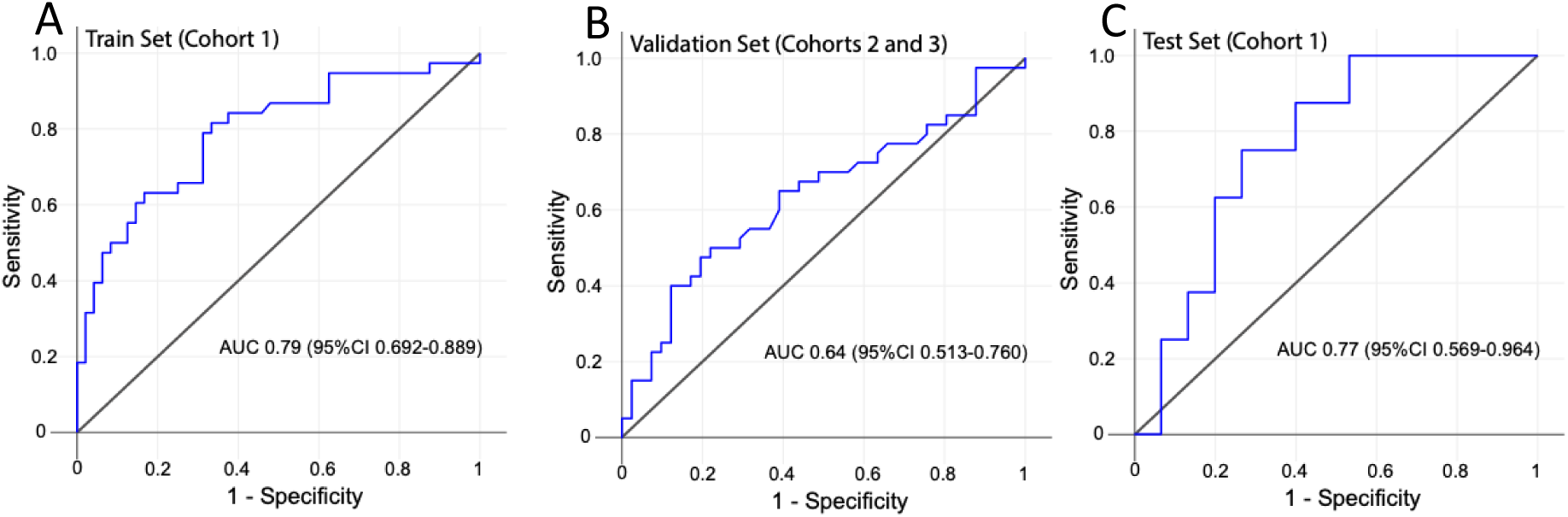
ROC curves for the AD-iCAP Model M1. Performance is shown for (A) training data using repeated, stratified 5-fold cross-validation (mean ROC across outer folds), (B) external validation (cohorts 2–3; n=82), and (C) the blind test set (cohort 1; n=23 ). AUC (95% CI) are shown on the graphs.

Model M1 used five features selected through a two-step reduction: initial filtering from 20,000 to 84 genes using independent mouse data, followed by correlation- and model-based reduction (correlation scores were <80% and feature importance scores were statistically higher than shadow features). This feature reduction approach generally yielded generalizable models in the model parameterization study in contrast with other feature reduction approaches or the control condition of no feature reduction, which appeared to be overfit (Fig. S5). Median expression patterns of the 5 genes across training, validation, and test sets showed general coherence, although most effect sizes were small and only a few comparisons were significant (Fig. 5), consistent with known pre-analytical variability and disease heterogeneity. These findings support the value of multivariate ensemble models like RF for overcoming pre-analytical variation and classifying complex, heterogeneous diseases like AD.

**Figure 5.**
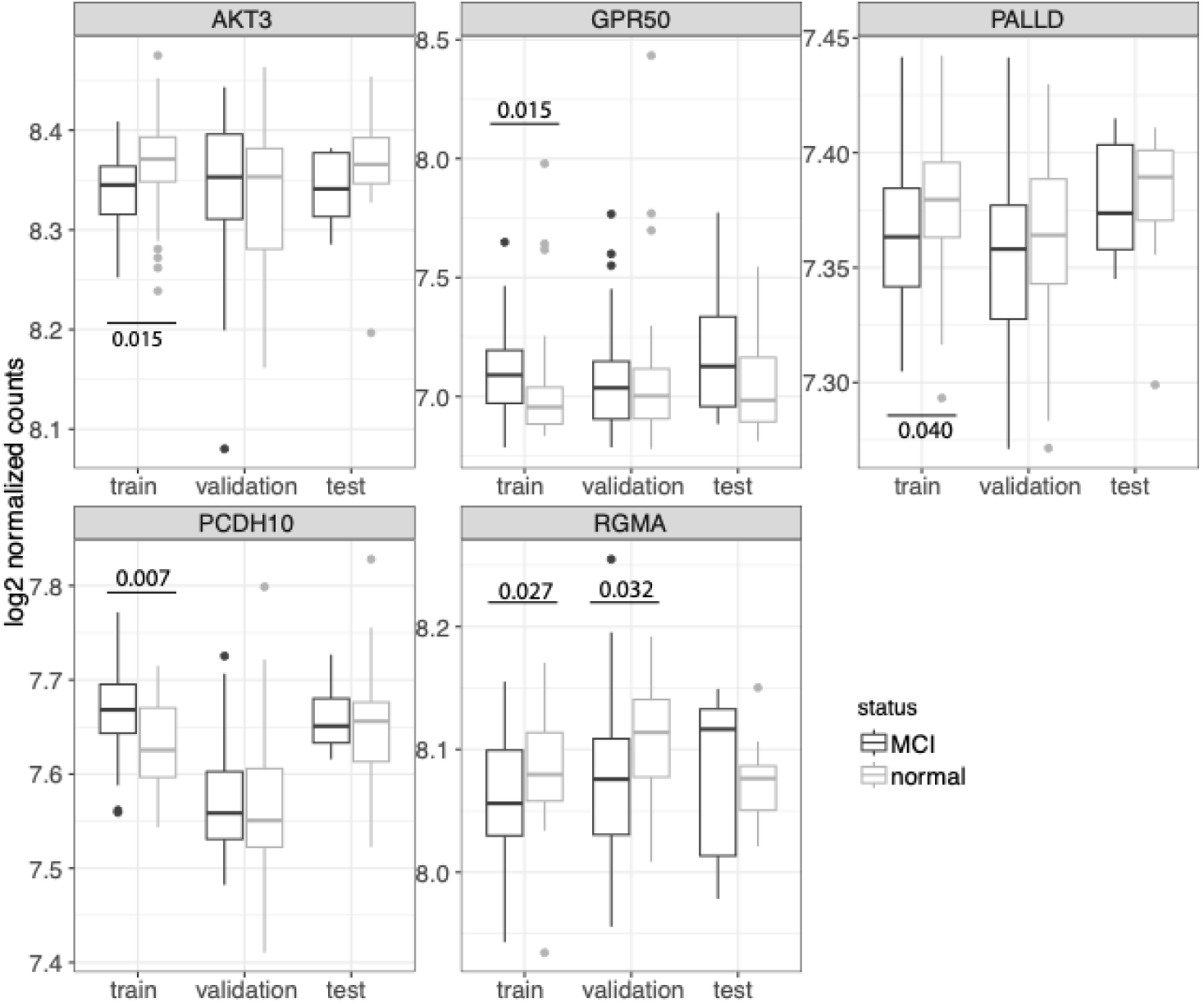
Box plots showing expression of five gene features in Model 1. Median, upper, and lower quartiles are shown across the training, validation, and test sets. False discovery rates (FDRs) are noted for comparisons with FDR < 0.05. Effect sizes (Cohen’s *d*) in the training set are -0.58, 0.44, -0.50, 0.80, and -0.53, respectively. The moderate effect sizes and interquartile spread highlight sample heterogeneity, underscoring the utility of ensemble modeling approaches. All features had similar importance scores in the model with mean decrease Gini values of 7.4, 9.2, 7.1, 9.3, and 8.9, respectively.

Note that two other models (M2 and M3) were trained, validated and tested on the blind set. They used the same approach as described here *except* model training, validation and testing used only cohort 1 samples. M1 was presented because its validation was more rigorous as it used samples from all 3 sources. On the blind set, M1 achieved AUC of 0.77 (95% CI: 0.57–0.96) with DeLong p = 0.00815. This remains significant under Holm control for the three models (α = 0.0167), but not after controlling family-wise error across all blind-set queries conducted historically (m = 15; α ≈ 0.0033). We therefore interpret the blind-set result as supportive, not confirmatory, pending prospective validation in an independent cohort.

#### Disease relevance

We took several approaches to investigate the AD relevance of the fixed feature sets and of the M1 classifier genes. First, we compared the mouse AD-iCAP expression profiles to the human ‘AD portrait’ profile, which is profile from a compendium of AD expression data from various brain regions and patients.^50^ We compared the case versus control AD-iCAP readout for each of the 9 conditions to the human AD versus normal profile by first separating the genes into previously defined consensus clusters perturbed in human AD brain regions^51^ and then measuring the correlation coefficients for each cluster.^47^ We found that mouse AD-iCAP data for the most advanced MCI stage (7 month old mice) had significant positive correlations with the human AD profile across multiple clusters (p-value < 0.05, Figs. 6, S6).

**Figure 6.**
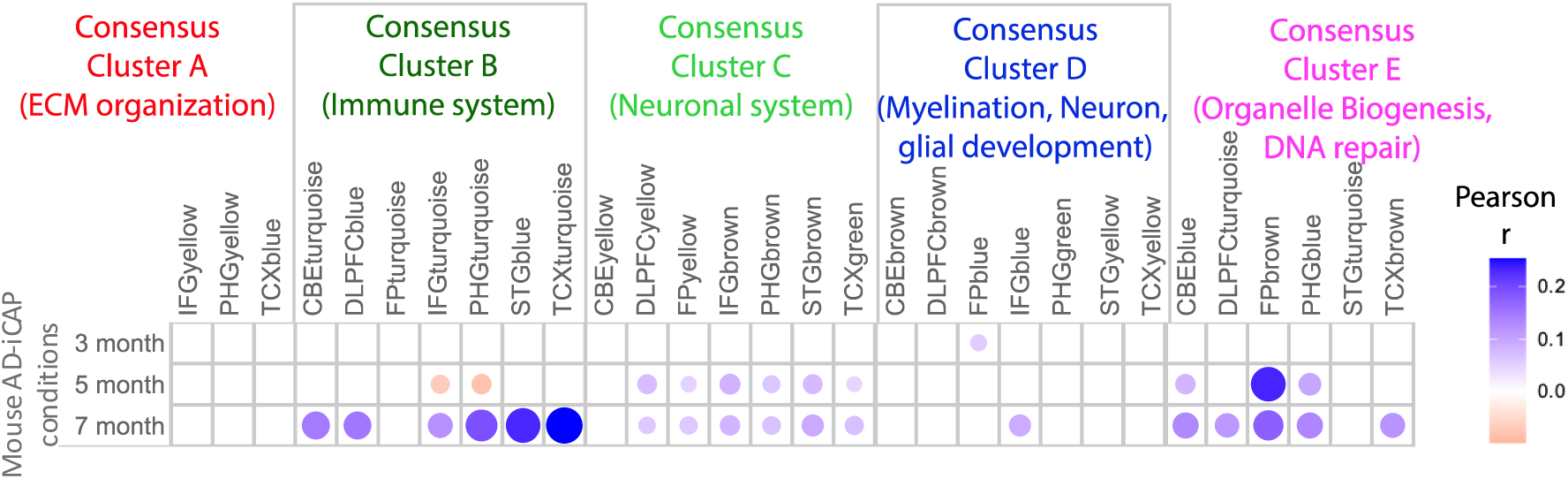
Heatmap showing mouse AD-iCAP gene expression profiles correlate with AD-associated expression patterns in human brain tissue. We first separated genes into co-expression modules defined by the Accelerating Medicines Partnership for Alzheimer’s Disease (AMP-AD) (column labels). Next, For each module, pairwise Pearson correlation coefficients were calculated between an AD expression profile from human brain tissue (‘AD portrait’ profile, Hill et al., 2022) and those observed in the AD-iCAP in response to mouse plasma at three mouse ages (row names). Blue and pink circles represent positive and negative correlations, respectively; color intensity and circle size reflect the magnitude of the correlation coefficient. Only correlations with *p* < 0.05 are displayed. A total of ∼12,000 genes are represented, which may belong to more than one module. Mouse and human profiles are case versus control differential expression. The mouse ages correspond to preAD (3 month) and MCI stages (5-7 months). Data for all 9 mouse experiments are shown in Figure S6.

Next, we analyzed AD-iCAP differentially expressed genes (Sets 1 and 2) using the STRING database.^52^ Both sets showed significantly greater network connectivity than expected by chance (p < 1.0e-16), suggesting functional coherence. Enriched Reactome pathways included cholesterol biosynthesis (SREBF/SREBP-activated), NTRK1 signaling (linked to cognitive decline via mitophagy), and BMP signaling (associated with APP/Tau-related deficits).^53–55^ To visualize pathway involvement, we mapped differentially expressed genes from 7-month-old mice onto the network (Fig. 7A; STRING network permalink https://version-12-0.string-db.org/cgi/network?networkId=bQ67JnOx9KVS) and overlaid expression data in Cytoscape (Fig. 7B).^56^ We observed consistent downregulation of cholesterol biosynthesis genes and varied patterns across other pathways.

**Figure 7.**
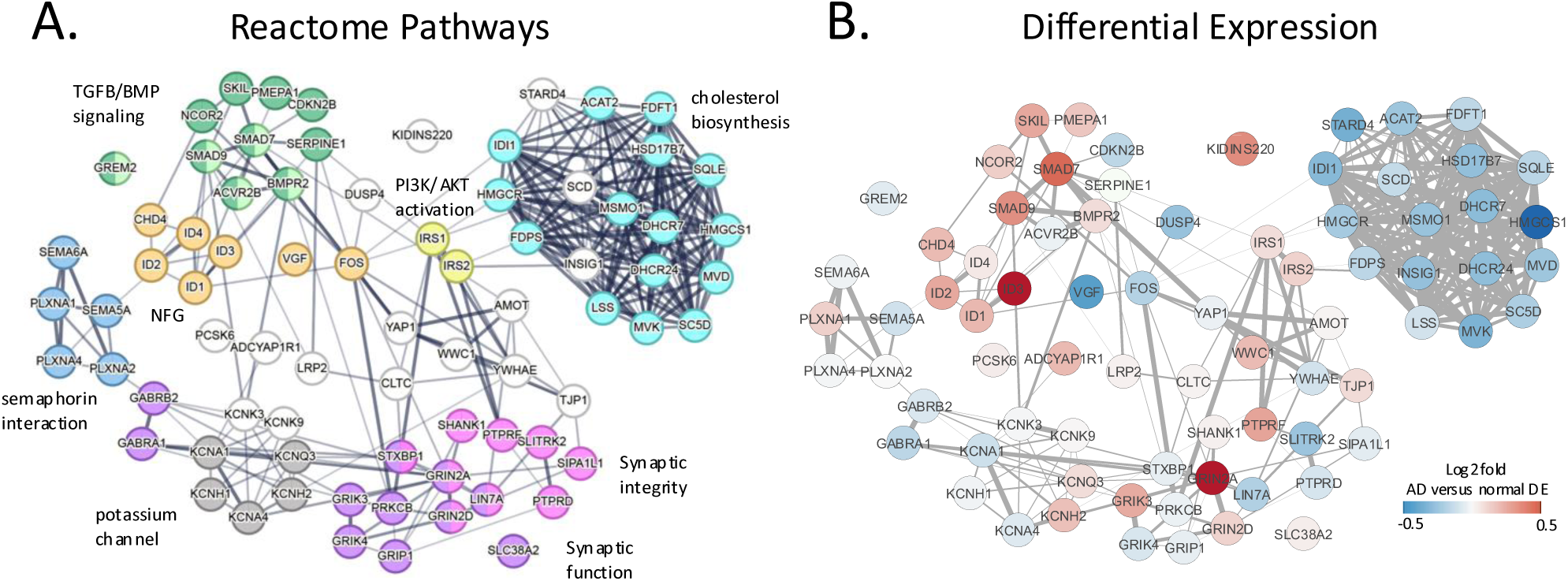
Network analysis of differentially expressed genes in the mouse AD-iCAP dataset. A protein interaction network was constructed using STRING v12.0 for 76 transcripts that were differentially expressed (DE) in at least one of the 9 mouse AD-iCAP conditions and mapped to significantly enriched Reactome pathways (FDR < 0.05; strength > 0.4). ***A*.** Nodes are colored by membership in 9 of the 44 enriched pathways: cyan, cholesterol biosynthesis; pink, protein-protein interactions at synapses; purple, transmission across chemical synapses; dark green, TGF-β signaling; light green, BMP signaling; orange, NGF-stimulated transcription; yellow, PI3K/AKT activation; gray, voltage-gated potassium channels; dark blue, other semaphorin interactions. Edge thickness represents confidence in functional or physical interaction. ***B*.** Differential expression values (AD vs normal) for the condition using plasma from 7-month-old mice on neurons precultured for 5 days. Expression values are shown on a two-color scale; 3 genes beyond the plotted range are shown using the darkest red (upregulated) or blue (downregulated). Comparison of panels A and B highlights pathway-specific coherence in expression, with pathways such as cholesterol biosynthesis and PI3K/AKT activation exhibiting consistent directionality.

As a final method to investigate the relevance of the mouse AD-iCAP data to AD, we compared the mouse AD-iCAP responses to iCAP responses to a pathogenic peptide of amyloid beta (oAB (25-35).^35^ To generate the amyloid-beta iCAP data, three iCAP experiments were performed in triplicate using 5-10 mM of peptide (oAB versus a scrambled, non-pathogenic version (sAB)) for 6-24 h. Robust differential expression was observed for each condition by NanoString analysis using the NeuroPathology gene panel of 770 genes. Next, pairwise Spearman correlations were calculated across all AD iCAP gene expression signatures (including all oAB experiments and mouse plasma experiments) for ∼550 genes measured in both experiments. As expected, significant positive correlations were observed within oAB experiments and within plasma experiments. In contrast, comparisons between plasma-exposed and oAB-exposed cells yield weaker correlations that were positive or negative depending on AD stage of the mice (Fig. S7).

Model M1 classifier genes were analyzed and four of the five genes have well-established relevance to AD: *AKT3* (*AKT serine/threonine kinase 3*), one of three *AKT* isoforms, is implicated in AD pathogenesis, as dysregulation of AKT signaling contributes to development of AD.^57^ The promoter of *GPR50* (*G protein-coupled receptor 50*) is specifically hypomethylated in Chinese Han males with AD.^58^ *PALLD (palladin, cytoskeletal associated protein)* ranks among the top 20 dysregulated “AD portrait” genes.^50^ *RGMA (repulsive guidance molecule A)* has multifaceted roles in neurodegeneration and accumulates in amyloid plaques in AD patients.^59^ None of the classifier genes met the threshold for membership in significantly enriched Reactome pathways in Figure 7, but *AKT3* and *GPR50* have well-established roles in PI3K/AKT activation^57^ and HDL-cholesterol levels,^60^ respectively.

Together, these results demonstrate that the AD-iCAP differential readout and features of the predictive model M1 reflect biologically relevant AD pathways and genes. Furthermore, our data suggest that the transcriptional responses of the indicator neurons to AD plasma are not simply explained by direct exposure to pathogenic amyloid-β in the plasma, but instead reflects a more complex cellular response to multiple analytes present in plasma.

## Discussion

AD is a heterogeneous neurodegenerative disorder with complex causes, beginning years before symptoms appear. These factors likely contribute to the limited efficacy of treatments and the high failure rate of clinical trials.^3^ Increasing evidence suggests that some therapeutics may have efficacy in specific AD subtypes, underscoring the need for early intervention and disease stratification.^3^

We developed the AD-iCAP, a next-generation cell-based diagnostic assay designed to advance precision medicine in AD and address the need for non-invasive, multivariate classifiers that detect early disease and enable patient stratification. Orthogonal to Aβ- and Tau-based blood tests, the AD-iCAP could be used in combination with these assays to improve diagnostic accuracy and enable targeted drug development.

The platform uses standardized neurons as biosensors to transduce weak, complex AD signals in blood into a multivariate cell-based gene expression readout, which is then modeled with machine learning. Using this approach, we optimized and validated the AD-iCAP to distinguish patients with MCI due to AD from unaffected individuals, achieving an AUC of 0.64 in an external validation set and an AUC of 0.77 in blind testing (Fig. 4).

A key advantage of the AD-iCAP is that it enables application of systems biology approaches to blood-based diagnostics. Detecting disease signals in blood is inherently challenging due to the low abundance and variability of analytes within a complex mixture from the entire body. Circulating biomarkers often lack a well-defined background population of analytes, making it difficult to normalize signals or apply many of the statistical and systems biology tools useful for cell-based assay development and for establishing disease relevance or biomarker relationships. The iCAP addresses these challenges by using cells, which are inherently “systems”, to enable expression normalization and application of multivariate analyses (e.g., GSEA, PCA, reference dataset correlations, network analyses) for assessing assay reproducibility, detecting and mitigating pre-analytical noise, and evaluating biological relevance.

After optimizing the assay to reduce technical noise (Figs. S1–S3), we observed that analytical variation was sufficiently low to support detection of biological signals in aggregate data across multiple batches of samples (Fig. S4B). However, PCA of the AD-iCAP patient data revealed low signal-to-noise, largely due to pre-analytical variation from patient heterogeneity and sample source effects (Fig. S4C). To address this, we developed an external feature selection approach using plasma from the 5XFAD mouse model of AD, stratified by disease stage. This approach minimized variability arising from disease stage, patient heterogeneity, and sample handling or storage compared to using study samples. Importantly, feature selection was independent of study samples and unbiased by prior AD biology, reducing the chances overfitting and enhancing generalizability.^61^

This approach yielded a concise fixed set of 84 genes for modeling that is amenable to targeted, high-throughput measurement platforms such as NanoString™ or QuantiGene™ simplifying follow-up studies and aligning with regulatory expectations for deployment. In addition, the panel is useful for developing the AD-iCAP for other applications such as unsupervised patient stratification methods called for in future AD drug development.^3^

Using the 84-gene feature set, we trained multiple models and selected Model M1 based on performance on a training set and on an external validation set comprised of two independent cohorts. M1 was then tested on a blind set and demonstrated AUC of 0.77 (95% CI 0.569-0.964) (Fig. 4). The final classifier was based on the expression of five genes (Fig. 5). The biological functions represented by these genes are well-documented in AD pathogenesis, providing mechanistic plausibility for the model’s predictive capacity.

We further assessed biological relevance in the mouse model readout through two complementary analyses: (1) molecular network analysis and pathway enrichment, which revealed processes implicated in AD pathogenesis, including cholesterol biosynthesis, synaptic structure and neurotransmission, PIK3/AKT activation and TGF-β signaling (Fig. 7); and (2) correlation with the human “AD portrait” transcriptome profile^50^, which was strongest in older mice at the MCI stage, consistent with the accumulation of AD molecular signatures over time (Figs. 6, S6). Notably, the AD-iCAP signal was distinct from that induced by direct exposure to amyloid-beta (Fig. S7), underscoring its ability to capture a broader, multivalent disease signature.

### Limitations and future directions

Our design included external feature selection, pre-specified pipelines, and independent testing, reducing the risk of type I error,^61,62^ and within this study, Model M1 showed statistically significant discrimination. Nevertheless, the historical reuse of the same blind test set for 12 prior evaluations increases the family-wise error on that dataset and may yield optimistic estimates of significance. In addition, site-to-site pre-analytical differences and extended storage times likely attenuated performance. Together, our results provide early supportive evidence of diagnostic potential and justify a prospectively designed, independent confirmation study in a larger, freshly collected cohort.

The AD-iCAP is a scalable, cost-effective diagnostic platform for early AD detection and patient stratification. Its independence from Aβ or Tau biomarkers makes it complementary to current assays and possibly useful for combinatorial use. Its multivariate, systems biology-based approach offers flexibility for adaptation to other applications, including prediction of therapeutic response. This platform has the potential to accelerate drug development, enhance clinical trial stratification, and ultimately improve patient outcomes. Future work will include validation in larger, prospective cohorts, evaluation of longitudinal performance, and assessment of assay scalability to ensure robustness and compliance with regulatory requirements for clinical deployment.

## Data Availability

All data produced in the present study are present in the paper and/or the Supplementary Materials, except RNA-seq data related to this paper will be available from GEO under the accession numbers after peer reviewed publication. R code for disease classification model M1 will be available at after peer reviewed publication.

## Abbreviations

Aβ: beta-amyloid
AD: Alzheimer’s disease
AUC: area under the ROC curve
CDR: clinical dementia rating
CI: confidence interval
CQN: conditional quantile normalization
CSF: cerebral spinal fluid
CV: coefficient of variation
FQN: full quantile normalization
iCAP: indicator cell assay platform
IRB: institutional review board
MCI: mild cognitive impairment
PCA: principal component analysis
PET: positron emission tomography
preAD: preclinical AD
ROC: receiver operator characteristic

## ACKNOWLEDGEMENTS

We gratefully acknowledge Anne M. Fagan, PhD, and the Knight Alzheimer Disease Research Center (ADRC), Washington University in St. Louis, for thoughtfully selecting and providing study samples and for their valuable advice throughout this work. Research reported in this publication was supported by the National Institute of Aging (NIA) and National Cancer Institute (NCI) of the National Institutes of Health (NIH) under Award Numbers R44AG051282, R43CA203455, R44CA203455 and R43AG056215. We are grateful to three studies for providing patient plasma samples and clinical data for this research: 1) The AIBL study (www.AIBL.csiro.au) is a collaboration between CSlRO, Edith Cowan University (ECU), The Florey Institute of Neuroscience and Mental Health (FINMH), National Ageing Research Institute (NARI) and Austin Health. It also involves support from CogState Ltd., Hollywood Private Hospital, and Sir Charles Gairdner Hospital. The study received funding support from CSIRO, the Science and Industry Endowment Fund (www.SlEF.org.au), NHMRC and Dementia Collaborative Research Centres (DCRC), Alzheimer’s Australia (AA), Alzheimer’s Association and the McCusker Alzheimer’s Research Foundation. 2) Knight Alzheimer’s Disease Research Center (ADRC) at Washington University in Saint Louis, supported by the NIA under Award Numbers P50AG005681 and P01AG03991, and P01AG026276. 3) University of Washington ADRC, supported by the NIA under award Number P50AG05136, Pacific Northwest Udall Center (P50NS062684) and Department of Veterans Affairs. We thank all the participants who took part in this study and the clinicians who referred participants. The content is solely the responsibility of the authors and does not necessarily represent the official views of the NIH.

## Conflicts of Interest

The funders provided support in the form of research materials and salaries but had no role in the study’s design, data collection, analysis, decision to publish, or manuscript preparation. JJS, RJL, JDB, MDD, YQ, and GAW are current or past employees of PreCyte Inc. Additionally, JJS, JDA, JDB, MDD, YQ, RJL, SAD, GAW, and LRM have equity interests in PreCyte Inc., which is developing products related to the research described in this publication. The commercial affiliation with PreCyte Inc. does not alter our adherence to journal policies on data and material sharing. All other authors declare no competing interests. EP, GL, CLM, and AF report no conflicts of interest.

## Notes

### Author Declarations

WCG IRB waived ethical approval for this work

