## Supplementary Figures for "An Indicator Cell Assay-based Multivariate Blood Test for Early Detection of Alzheimer’s Disease"

A

| | 2 d | 5 d |
|---|---|---|
| Mean intra-class CV (CPM) | 9.18 | 8.52 |
| number of DEGs (p-value < 0.05) | 1835 | 646 |
| Mean absolute log2 fold change | 0.078 | 0.067 |

Genes with significant differential expression in either condition were compared between 2d and 5d and had significant Pearson correlation.

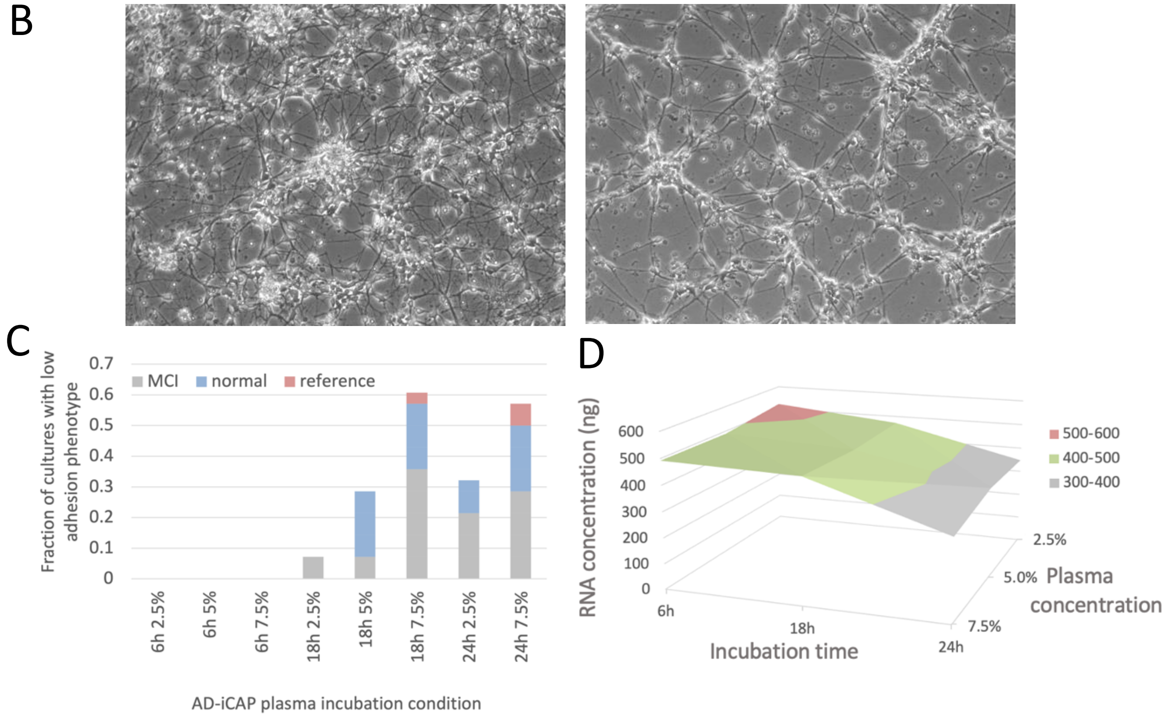


**Figure S1. Optimization of AD-iCAP experimental conditions. A.** Neuron pre-culture time was optimized using technical replicates of 20 plasma samples (10 normal, 10 MCI). Neurons were pre-cultured for either 2 or 5 days, followed by 24 h incubation with 5% plasma. Two-day pre-culture yielded more differentially expressed genes (DEGs) with only a modest increase in variability, supporting its selection for subsequent experiments. **B.** A sporadic, condition-dependent neuronal phenotype was observed in the AD-iCAP. Representative images show typical morphology (*left*) versus reduced adhesion and neurite density (*right*) in neurons exposed to 2.5% plasma for 24 h. The phenotype was likely driven by heparin added with the plasma, as similar effects were observed in heparin-only controls (not shown). **C.** The frequency of this phenotype was quantified across three incubation times and two plasma concentrations using 16 plasma samples per condition (6 MCI, 8 normal, 2 reference). Lower plasma concentrations and shorter incubation times reduced phenotype occurrence. **D.** A 3D plot illustrates RNA yield variability under the same conditions. Shorter incubation times resulted in higher RNA yield. Correlations were observed between phenotype frequency and time ($r = 0.43$), phenotype frequency and concentration ($r = 0.25$), and RNA yield and time ($r = -0.4$). Additional analysis (not shown) indicated that reducing plasma incubation from 24 to 6 h increased both the number of DEGs and the mean absolute $\log_2$ fold change. These data informed selection of final AD-iCAP parameters: 2-day neuron pre-culture followed by 6 h incubation with 5% plasma.

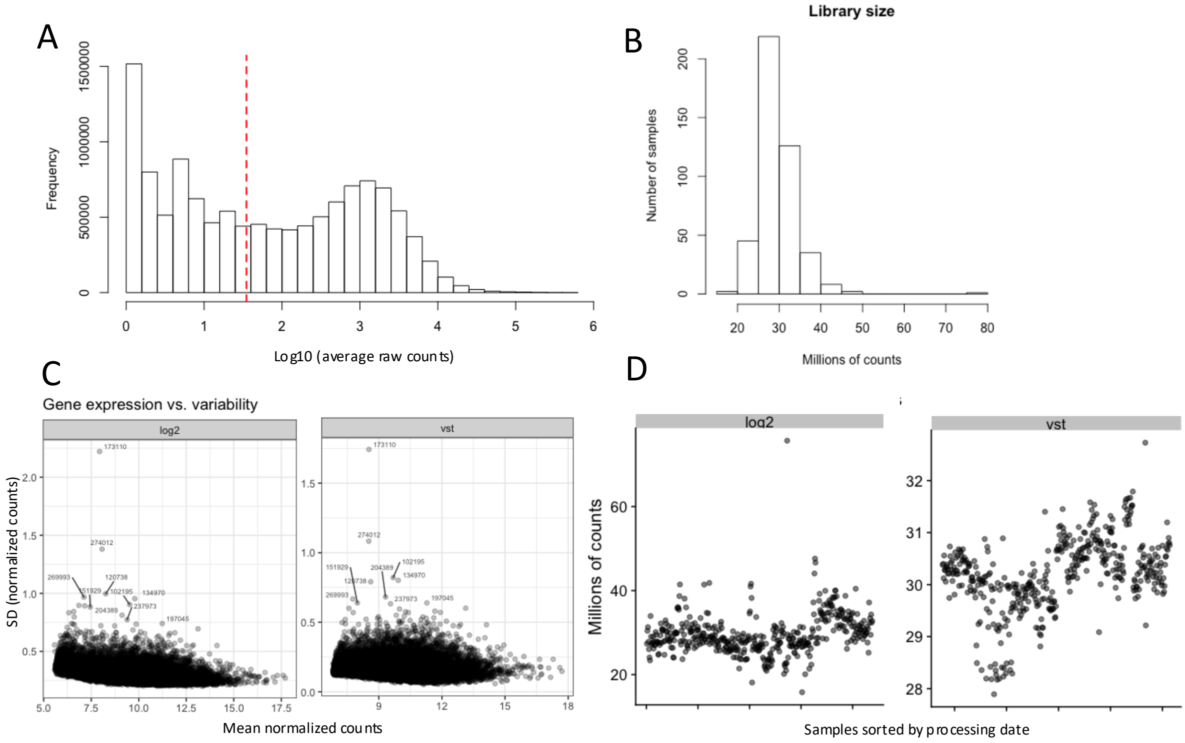


**Figure S2. iCAP RNAseq metadata preprocessing.** ***A.*** A histogram of $\log_{10}$ (average raw counts) for all genes in the AD-iCAP dataset showing bimodal distribution. The filter threshold for limit of detection is shown by a red dashed line corresponding to a lower limit of detection (LLD) of genes with 35 counts. ***B.*** A histogram of library sizes in millions of counts after removal of genes below LLD in at least 10% of samples. Total library size was approximately normally distributed with a mean around 27.5 million counts. ***C and D.*** Application of Variance Stabilizing Transformation (VST) to RNA-seq data reduced variability of gene expression measured by plotting standard deviation (SD) of each gene across all samples and by plotting raw count data for all samples with and without VST transformation.

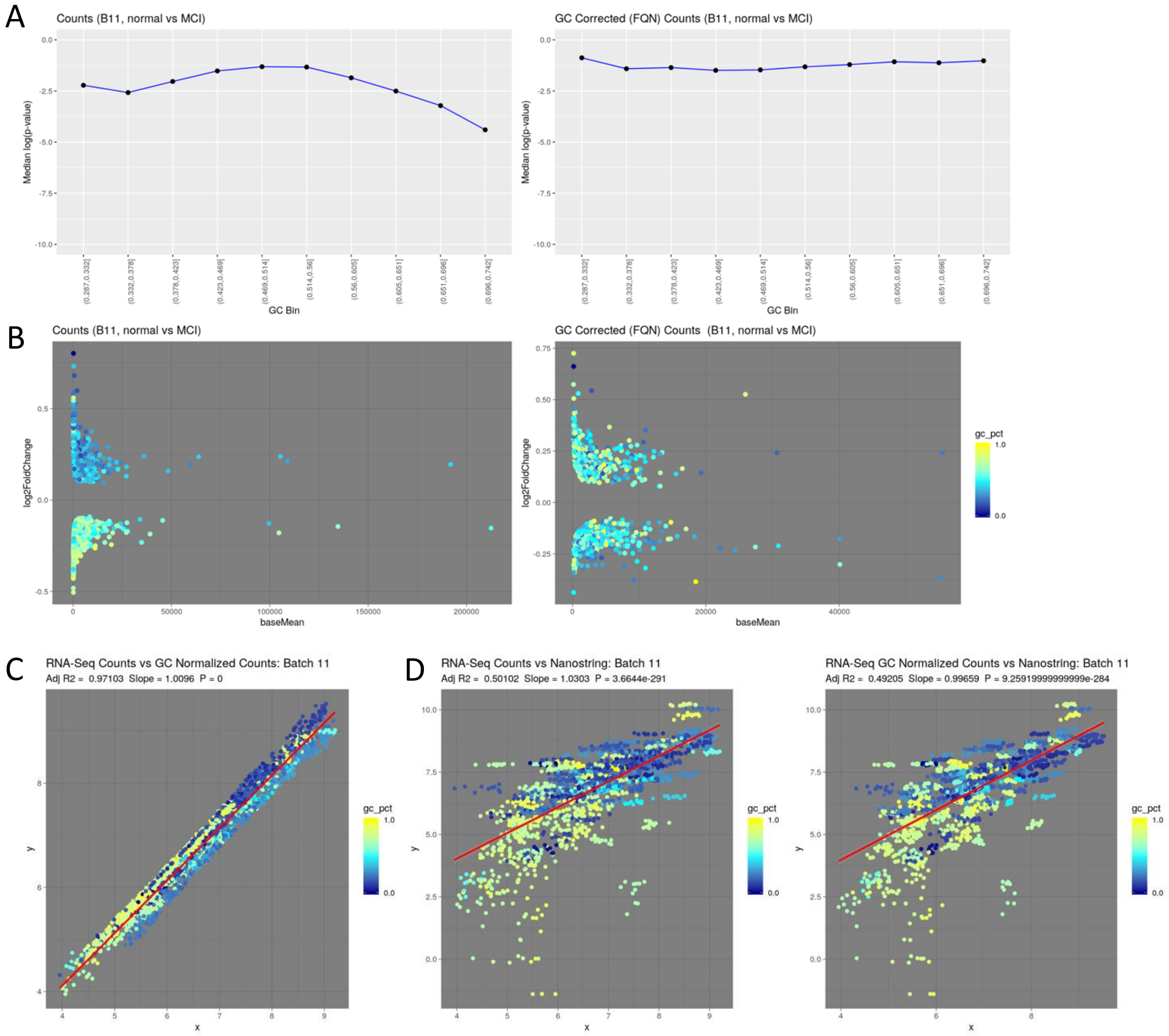


**Figure S3**. **GC bias correction of AD-iCAP RNA-seq data. A.** Analysis of the effect of GC content on significance of differential expression. Bias was observed before but not after GC bias correction with FQN. **B.** Analysis of the GC content of up- and down-regulated genes. Bias was detected before but not after FQN correction. Genes plotted in panels A and B were those with significant differential expression between MCI and normal samples in AD-iCAP batch 11 (FDR < 0.05). Bias was also detected in other AD-iCAP batches (not shown). **C.** Plot of gene expression levels (counts) before versus after GC bias correction**.** GC content appears to be correlated with gene expression levels and GC bias correction does not have large effects on gene rank. **D.** Comparison of AD-iCAP RNA-seq data and AD-iCAP NanoString data before and after GC bias correction. AD-iCAP RNA samples used for RNA-seq (5 preAD and 5 normal samples from batch 11) were analyzed by Nanostring using probes for 182 genes to compare expression across the platforms. Significant correlation of expression was measured across Nanostring (y-axis) and RNA-seq (x-axis) platforms (with and without GC bias correction applied to RNA-seq data), but R2 was moderate. This is likely due to low correlation for genes below the limit of detection for either platform (including genes with $log_2$ expression values below 5 in RNA-seq, corresponding to our previously defined lower limit of detection of 35 counts. *gc_pct*, GC percent where 1.0 - 100%.

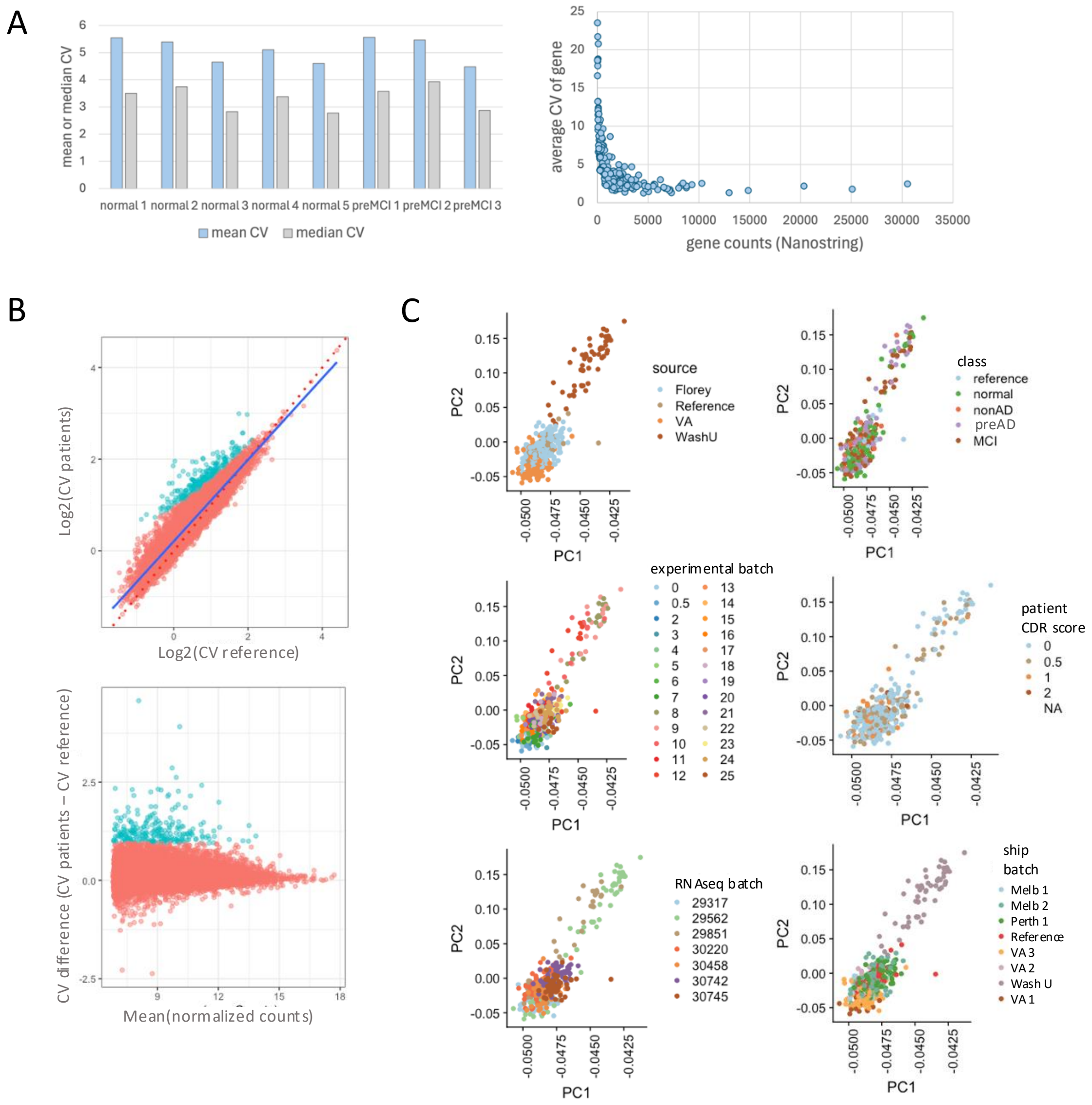


**Figure S4. Analytical variation in AD-iCAP gene expression data. A.** Within-batch variation was assessed using 3 technical AD-iCAP replicates from each of 8 patient samples measured by NanoString. Median coefficient of variation (CV) across 182 genes was low (~3.3%) for each sample (left). Mean CVs were slightly higher, driven by a subset of genes with very low expression levels (right). **B.** Between-batch variation was evaluated across the large-scale AD-iCAP RNA-seq dataset by comparing CVs across patient samples versus CVs across concurrently processed technical replicates of plasma controls. As expected, biological variation (patient CVs) was greater than technical variation (reference CVs), indicating that true biological differences were detectable above assay noise. A subset of 250 genes with high biological and low technical variation (blue nodes) was selected for further analysis. **C.** Principal component analysis (PCA) of the 250 selected genes, stratified by clinical and technical factors, showed that sample source contributed the greatest variance in the dataset. No clear separation by diagnostic class was observed, suggesting that class-specific expression differences are subtle and require multivariate modeling approaches. In panels B and C, genes with <35 counts were excluded, and data were variance-stabilizing transformed (VST).

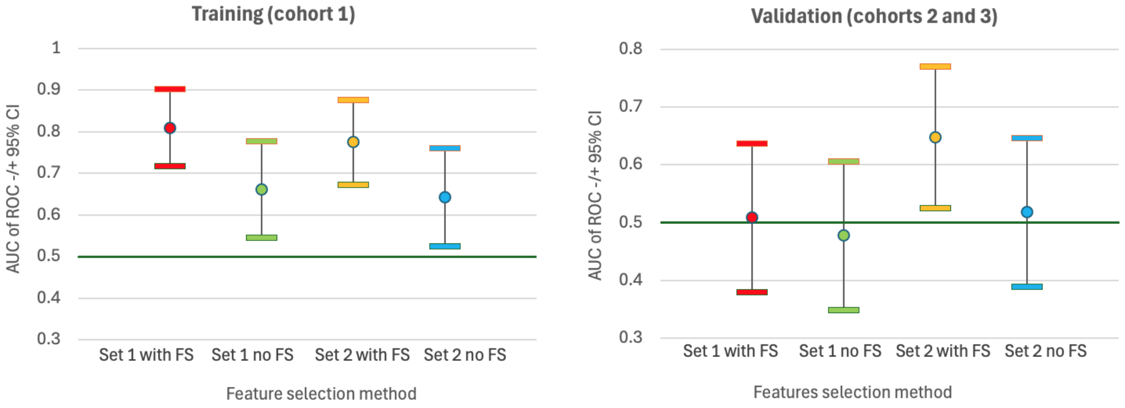


**Figure S5. Results of large-scale modeling study.** Average performance of all models trained using AD-iCAP features comparing MCI versus normal classes. Samples were binned by feature selection method. As expected, those without further feature reduction (no FS) did not have generalizable performance, likely due to overparameterization (with 84 or 398 features). Each graph is an average of 6 – 10 unique models including all models that converged. Model 1 used Set 2 with feature reduction.

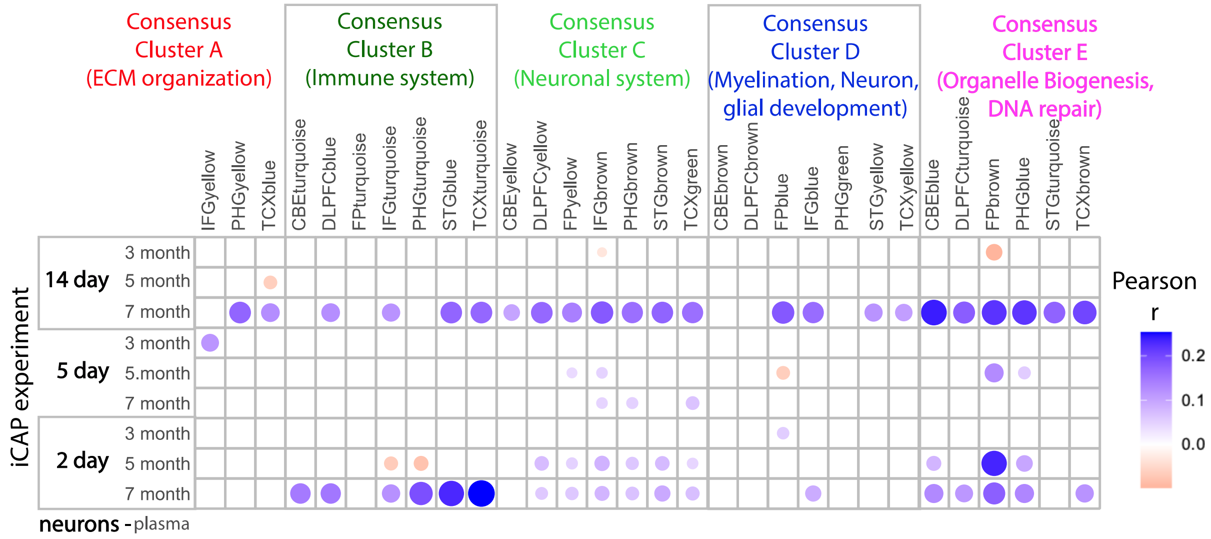


**Figure S6. Heatmap showing mouse AD-iCAP gene expression profiles correlate with AD-associated brain expression patterns.** We first separated genes into co-expression modules defined by the Accelerating Medicines Partnership for Alzheimer's Disease (AMP-AD) (column labels). Next, For each module, pairwise Pearson correlation coefficients were calculated between an AD expression profiles from human brain tissue ('AD portrait' profile, Hill et al., 2022) and those observed in the AD-iCAP in response to mouse plasma. The 9 experimental conditions used to generate the AD-iCAP data are shown at the left including neuron pre-culture times (days) and mouse ages (months). Blue and red circles represent positive and negative correlations, respectively; color intensity and circle size reflect the magnitude of the correlation coefficient. Only correlations with $p < 0.05$ are displayed. A total of ~12,000 genes are represented, which may belong to more than one module. Mouse and human profiles are case versus control differential expression.

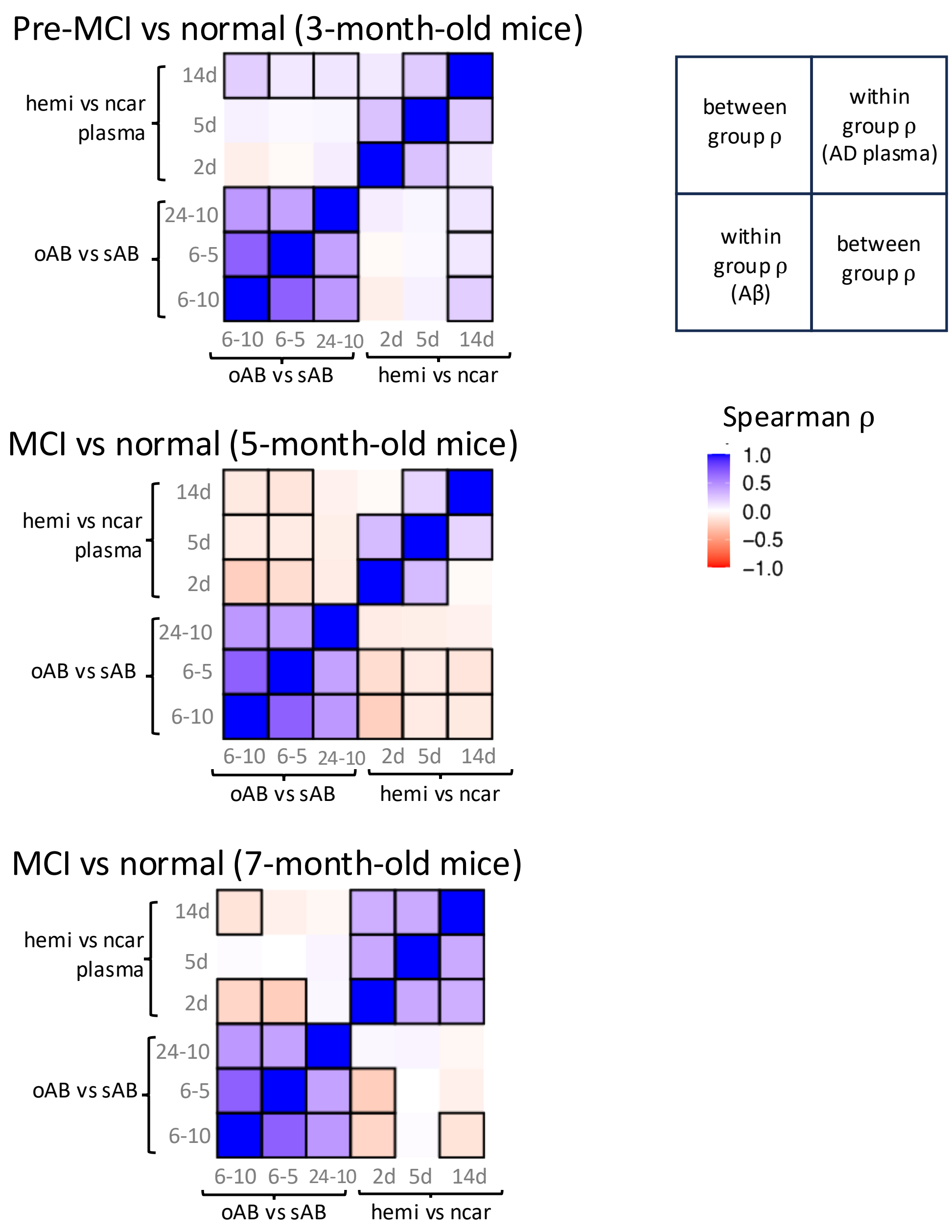


**Figure S7. Pairwise correlations of AD-iCAP gene expression signatures across experimental conditions.** Matrices show Spearman correlation coefficients (ρ) between six conditions assayed in the iCAP, using ~500 genes from the NanoString NeuroPathology panel (Hs v1.0). The three *hemi* versus *ncar* plasma exposures from the AD mouse model exhibit strong positive correlations with each other, and the three pathogenic versus non-pathogenic amyloid-β peptide exposures (oAB vs sAB) also correlate positively within their group. In contrast, comparisons between plasma-exposed and oAB/sAB-exposed cells yield weaker correlations, which are positive or negative depending on AD stage of the mice. This pattern suggests that the transcriptional response to plasma is not simply explained by direct exposure to pathogenic amyloid-β, but instead reflects a more complex cellular response to multiple analytes present in blood. Hemi vs ncar exposures were for 6 h using neurons that were precultured for 14, 5, or 2 days as indicated. oAB vs sAB exposures were for 6 or 24 h at concentrations of 5 or 10 µM as indicated using neurons precultured for 14 days. Significant correlations (FDR < 0.05) are outlined in black. oAB, pathogenic oligomerized Aβ peptide (aa 25-35). sAB, non-pathogenic scrambled peptide (scrambled aa 25-35). Data used to make this figure are in Data file 2.
